# The integrity of the antimicrobial supply chain in Bangladesh: assessing the regulatory environment and contextual challenges

**DOI:** 10.1101/2021.10.28.21265605

**Authors:** Ebiowei S.F. Orubu, Mohammed A. Samad, Md. Tanvir Rahman, Muhammad H. Zaman, Veronika J. Wirtz

## Abstract

Poor-quality medicines lead to large individual, health systems and national economic losses. Most low- and middle-income countries (LMICs) lack the robust regulatory quality assurance system necessary to contain the spread of poor-quality medicines. Innovations in regulatory systems strengthening are needed to protect public health. In this study, we assessed the integrity of the antimicrobial supply chain in Bangladesh and suggest innovations. We employed a qualitative methodology comprising a policy content analysis, and literature and database reviews. Bangladesh was chosen as a model LMIC because of its pharmaceuticals production capacity. Using a framework modified from the World Health Organization’s and the United States Pharmacopoeia’s, the Bangladesh National Drug Policy (BNDP) was evaluated for provisions on quality assurance mechanisms that match regulatory functions. Newspaper, peer-reviewed and post-marketing surveillance reports were used to assess the extent of poor-quality antimicrobials in circulation. There are provisions for quality assurance in the BNDP. Newspaper reports identified the circulation of substandard antimicrobials and reported on actions by the government to seize fraudulent/falsified and expired products. Only six peer-review studies testing antimicrobial product quality were identified; the three studies with the larger sample size (over 10 samples tested) reported results of out-of-specifications products. This assessment found evidence of regulatory actions against poor-quality antimicrobials as well as news reports and studies suggesting quality concerns, but no current rigorous assessment of the extent in Bangladesh. We suggest a multi-pronged approach to regulatory system strengthening comprising three strategies: community-based surveillance, task-shifting, and technology-enabled consumer participation.

## Introduction

Maintaining the integrity of the pharmaceutical supply chain to prevent the entry of poor-quality medicines continues to be a regulatory challenge for many countries. In 1995, a report of fatal diethylene glycol poisoning following the administration of contaminated paracetamol syrup to children emerged from Bangladesh (1). This case echoed similar incidents in the late 1930s in the USA, contemporaneously in Nigeria and would go on to recur in other countries, most recently in February 2020 in India (2–6). In 2017, the global prevalence of Substandard and Falsified (SF) medicines was estimated at 10.5%; based on data from mostly Low- and Middle-Income Countries (LMICs) in Africa. In April 2020, supply chain disruptions induced by the recent COVID-19 led to a rise in the incidence of falsified chloroquine, one of the medicines with putative efficacy against SARS-Cov2, in the World Health Organization (WHO) regions of Africa and Europe. These examples illustrate the persistence and prevalence of the challenge to pharmaceutical supply chain integrity, especially in LMICs, and the need for effective, proactive, and continuous regulatory oversight to protect public health. For LMICs with capacity constraints, there is the need for innovative approaches to regulatory systems strengthening.

The WHO defines poor-quality medicines as those that are SF. A substandard medicine is one that is out-of-specification, or fails to meet quality checks, while a falsified medicine is one that deliberately misleads as to content, or manufacturer (7). The impacts of poor-quality medicines extend beyond the individual to health systems and economies (8). Poor-quality antimicrobials contribute to antimicrobial resistance (AMR) with transnational impacts, reinforcing the need for stringent regulatory controls to ensure product integrity in the supply chain (9,10).

In many countries, the duty of ensuring the quality of medicines in the supply chain rests with the national medicines regulatory authority (NMRA), which is a public body with the technical and constitutional authority to provide an oversight function. However, most LMICs have limited regulatory capacity to perform all the functions necessary to protect the medicine supply chain against the entry of SF medicines (11). Current approaches to regulatory systems strengthening focus on improving capacity at the NMRA. However, the persistence of the problem of SF medicines in LMICs calls for further innovations in regulatory medicine quality assurance.

The aim of this study, as part of a broader Social Innovation on Drug Resistance program, was to assess the integrity of the antimicrobial supply chain in Bangladesh to understand the regulatory environment and contextual challenges. Understanding local contexts including the prevalence of SF medicines and gaps in regulatory approaches is important to any initiative to address challenges (12). While the WHO report gives a global prevalence of SF medicines, there are scant reports from other WHO geographical regions, including the WHO Southeast Asian region. Bangladesh, located in the WHO Southeast, has a large human and animal population and was selected as a model because of its pharmaceutical production capacity. The objectives were to: (i) evaluate the regulatory policies, and other mechanisms, for assuring the quality of antimicrobials in Bangladesh; (ii) identify any reports or publications on the prevalence of SF medicines.

## Methods

The study employed a qualitative methodology comprising document, literature, and database reviews to assess regulatory policies and any quality issues in the antimicrobial supply chain in Bangladesh.

### Policies

We performed a document review by searching the website of the NMRA, the Directorate General of Drug Administration (DGDA), for policies and regulations governing the pharmaceutical supply chain in March 2020 (13).

To evaluate the policy, we modified the regulatory functions frameworks of the WHO and the United States Pharmacopoeia, USP (14,15) to construct a framework covering pre-marketing authorization to post-marketing surveillance mechanisms for oversight of the five tiers of the pharmaceutical supply chain (Table 1). This framework grouped regulatory mechanisms into four: current Good Manufacturing Practice (cGMP); Good Distribution Practices (GDP); Good Pharmacy Practice (GPP) guidelines; and pharmacovigilance. The four mechanisms comprised 10 functions.

**Table 1.**
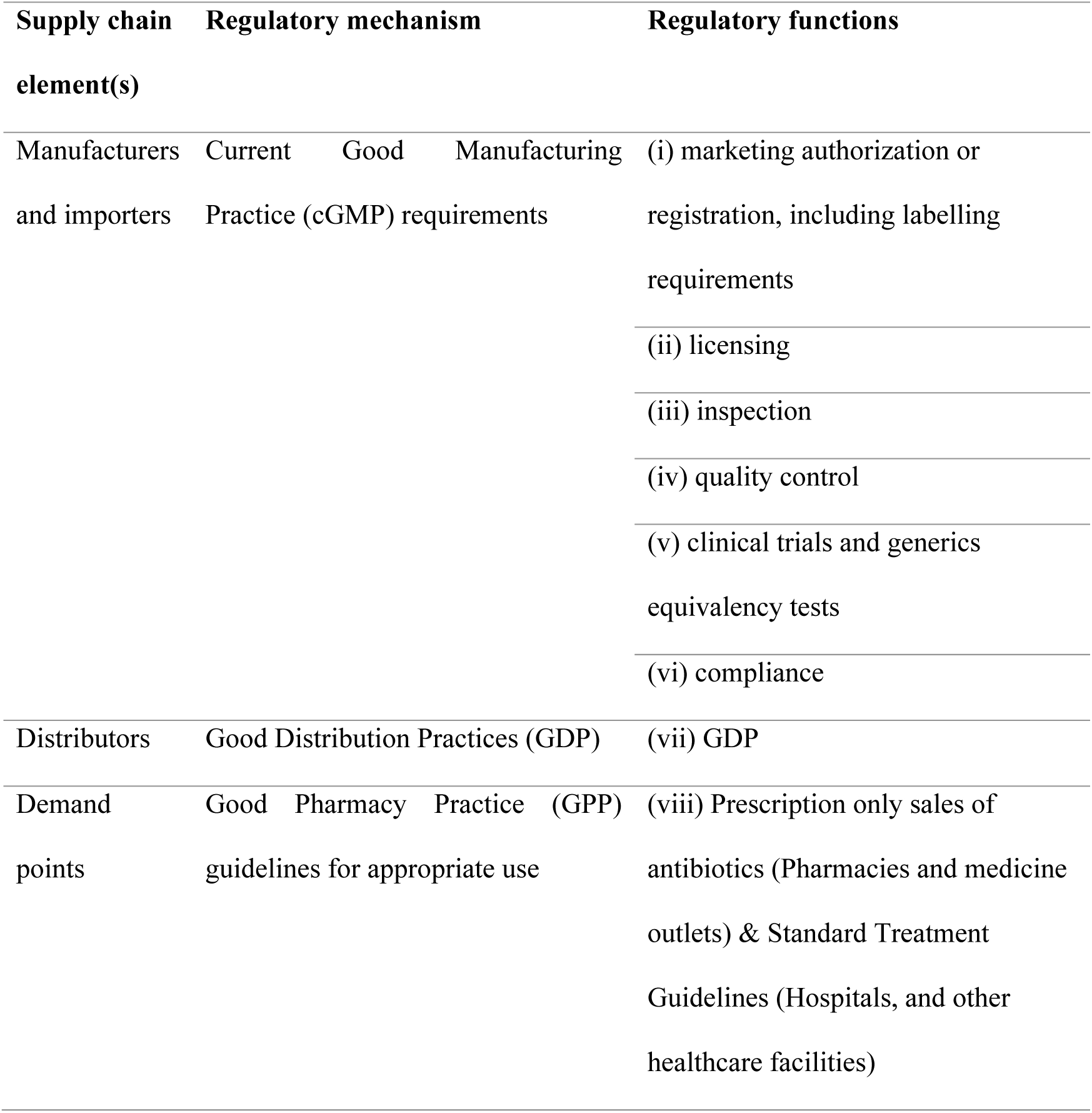
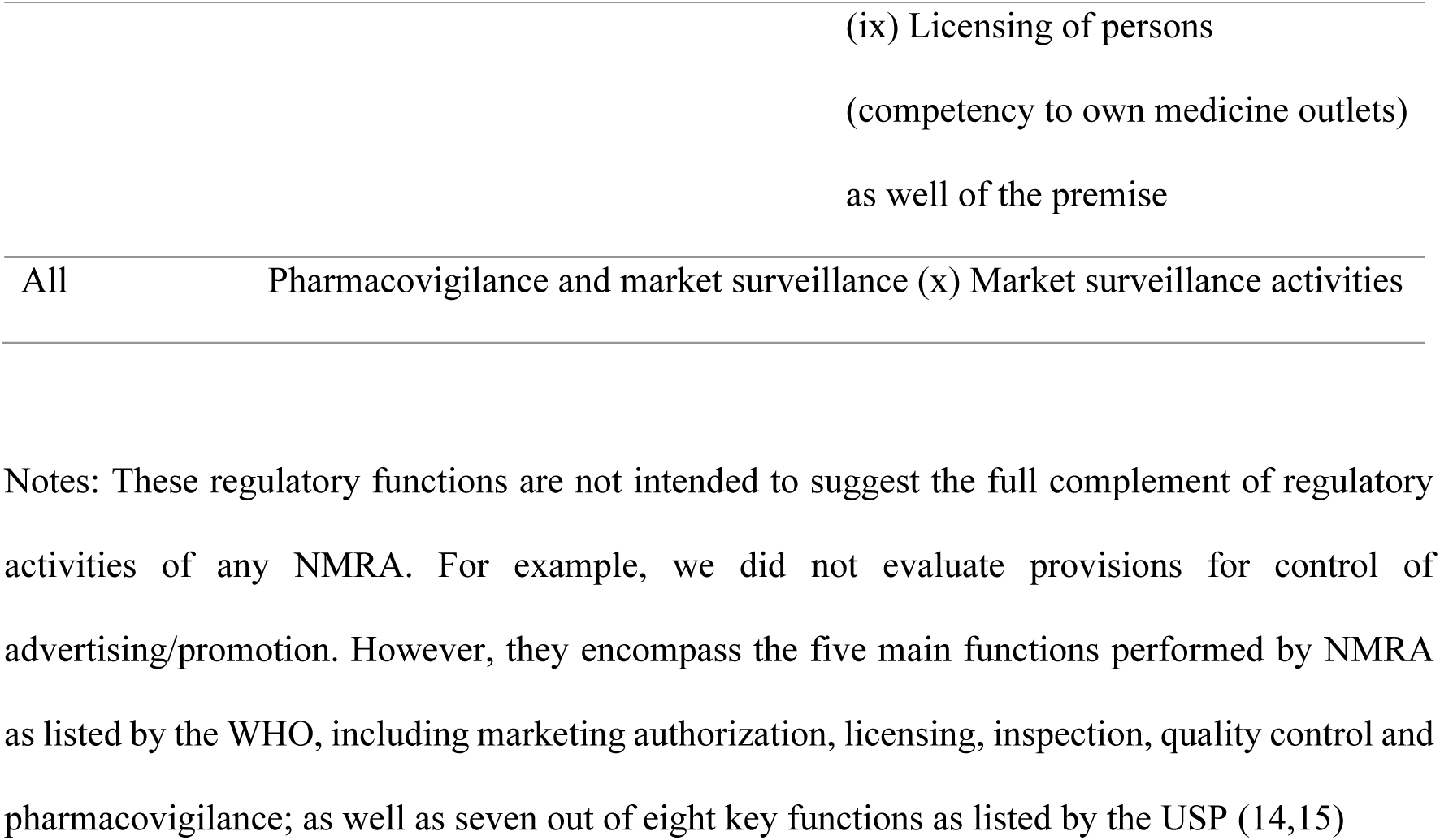
Framework for assessing the regulatory policy document of Bangladesh as adapted from WHO and USP, grouping 10 regulatory functions under four regulatory mechanisms covering all supply chain elements (14,15).

The relevant sections of the policy document corresponding to these grouped functions were identified and described. Additionally, any other quality assurance mechanism specific for antimicrobials identified from practice experience or knowledge were noted and described.

Market surveillance and quality issues: To map reports and publications on SF medicines and market surveillance activities, we conducted a combined literature and database review as detailed:

1. Literature review for reports on SF medicines: To identify media reports of SF medicines in circulation/supply chain and/or market surveillance, we performed a literature search for grey literature (non-peer reviewed newspaper reports). Literature from 2017-2020 were reviewed. The literature search was conducted twice in December 2018 and April 2020. The search strategy and extracted data are described in the Appendix.
2. Publications on quality testing: A review of published literature was performed on *Banglajol* (an online service providing access to Bangladeshi Journals), EMBASE and Google Scholar to identify reports of quality evaluations of antimicrobials in circulation in Bangladesh. The literature search was conducted on May 18-19, 2020, as detailed in the Appendix.
3. Database reviews for post-marketing surveillance reports: Three databases – the United States Pharmacopoeia, USP, Medicine Quality Database (MQDB) (16), the Oxford’s Infectious Disease Data Observatory’s Medicine Quality Monitoring Globe database (MQMG) (17), and the DGDA database (18) – were searched for any records of quality issues with marketed antimicrobials in Bangladesh. Searches were conducted in May 2020. For the first two databases, a keyword search was performed using “Bangladesh”. Any report of a poor-quality medicine was noted. For the DGDA database, reports of any post-marketing surveillance were retrieved.

## Results

These results present a composite view of the regulatory environment (policies and post-marketing surveillance) and media and peer-reviewed reports and publications respectively on the quality of antimicrobials in the pharmaceutical supply chain in Bangladesh.

### Policies

Four documents laying out government’s policies for ensuring medicine quality in Bangladesh were identified as:

- Drug (Control) Ordinance 1982.
- The Drug (Control) (Amendment) Ordinance 1984.
- Drug Control Ordinance 2006 and
- National Drug Policy 2016 (13).

The National Drug Policy 2016 (NDP) is the current document guiding quality assurance throughout the supply chain, and builds upon previous versions starting from 1982, the first in independent Bangladesh. It codifies both the protectionism and supply chain medicine quality assurance mechanisms for Active Pharmaceutical Ingredients (API) and Finished Pharmaceutical Products (FPP) for use in human and animal health in Bangladesh and is aligned with the National Health Policy of 2011. This document recognizes poor-quality medicines as those that are: fake, adulterated, expired, unregistered, counterfeit, misbranded, smuggled, and substandard.

We identified 10 mechanisms for the quality assurance of medicines in Bangladesh in the NDP covering all four pre-defined areas as well as a new provision for environmental protection against antimicrobial run-offs from manufacturing plants (Table 2).

**Table 2.**
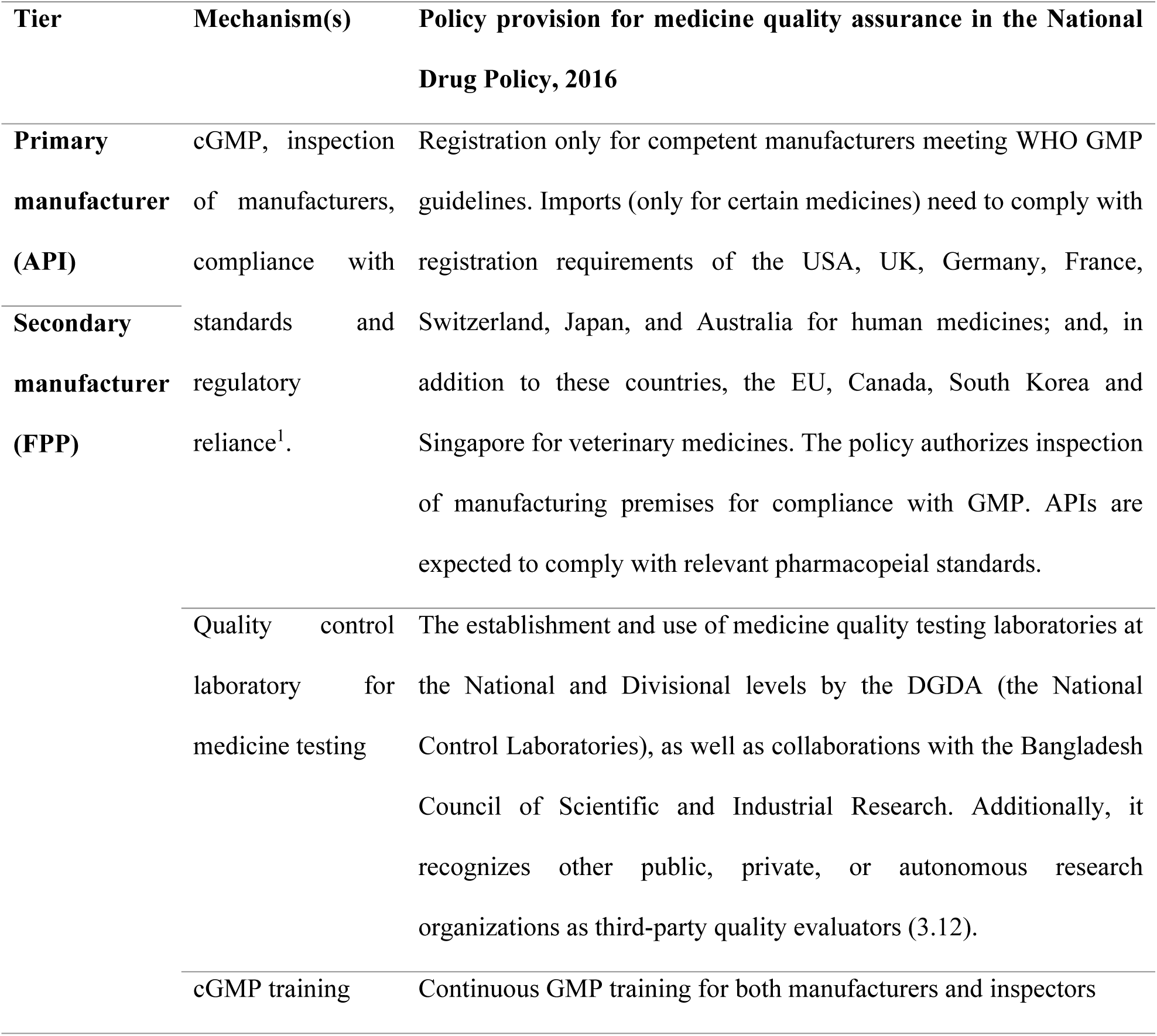
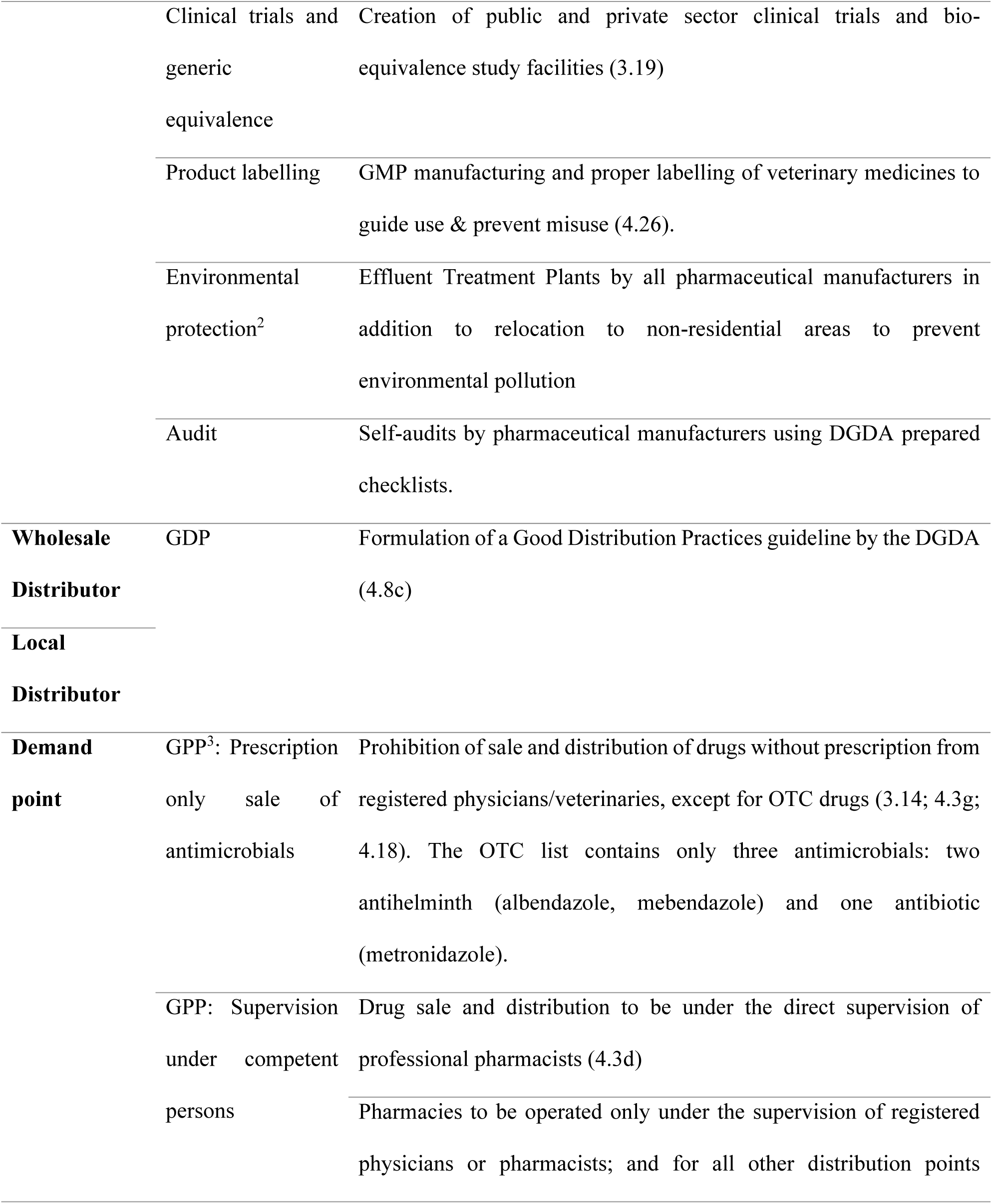
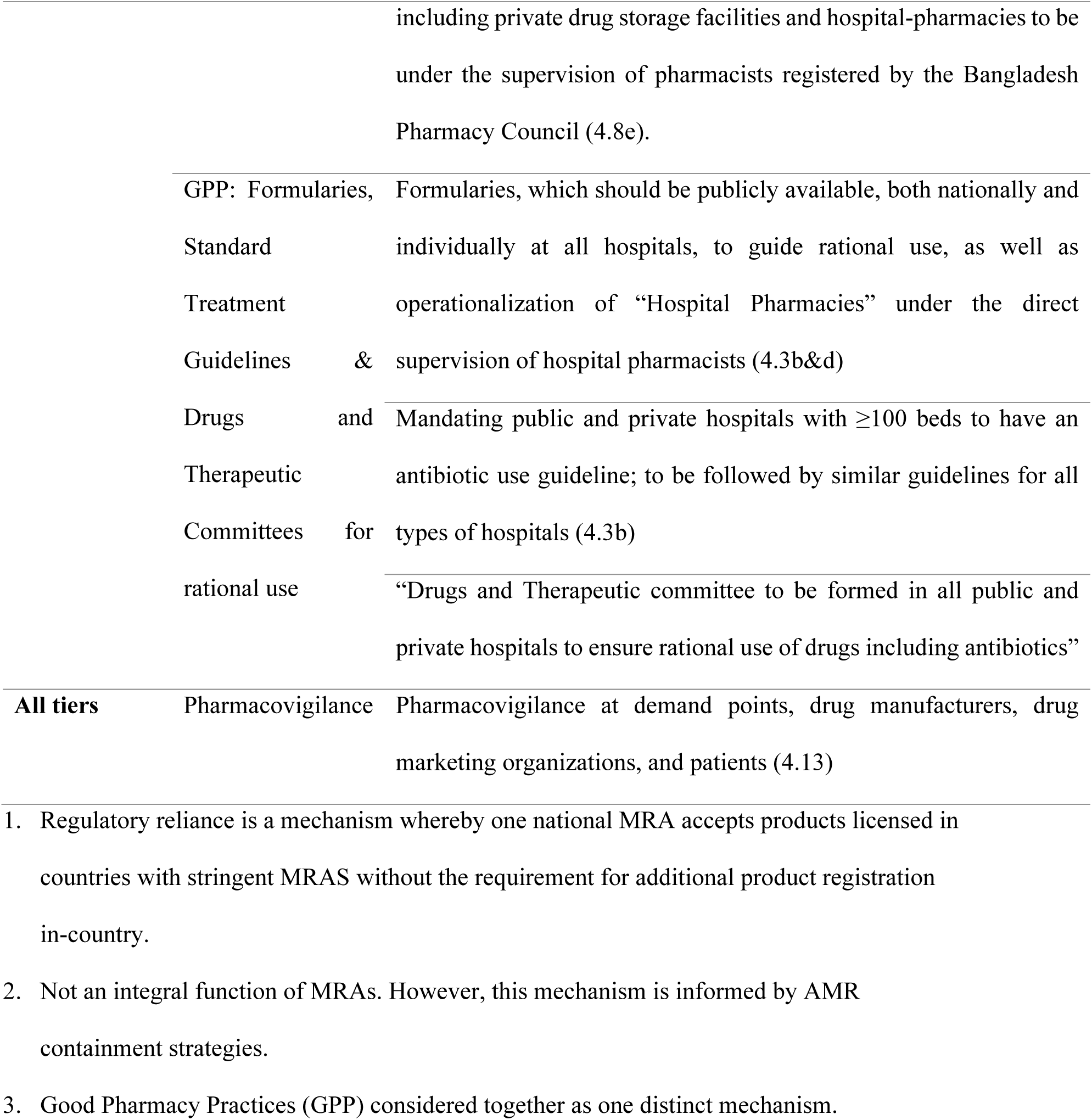
Policy provision of selected relevant regulatory mechanisms for the five tiers or elements in the antimicrobial/medicine supply chain. (Numbers in brackets are the exact sections of the BNDP.)

### Market surveillance and quality issues

#### Media reports

Out of 200 listed reports in The Daily Star newspaper of Bangladesh, 27 met the inclusion criteria. Of these, 12 reports were on SF medicines. In three of these reports, antimicrobials were expressly mentioned, and these were evaluated (Table 3). Two of evaluated reports were on falsified antibiotics in private facilities, which were seized, and the perpetuators arrested and/or fined (19,20). In one case up to seven different antibiotics were falsified, with most (57%, 4/7) being cephalosporins (20). The third was on expired anthelmintic distributed to a public facility.

**Table 3.**
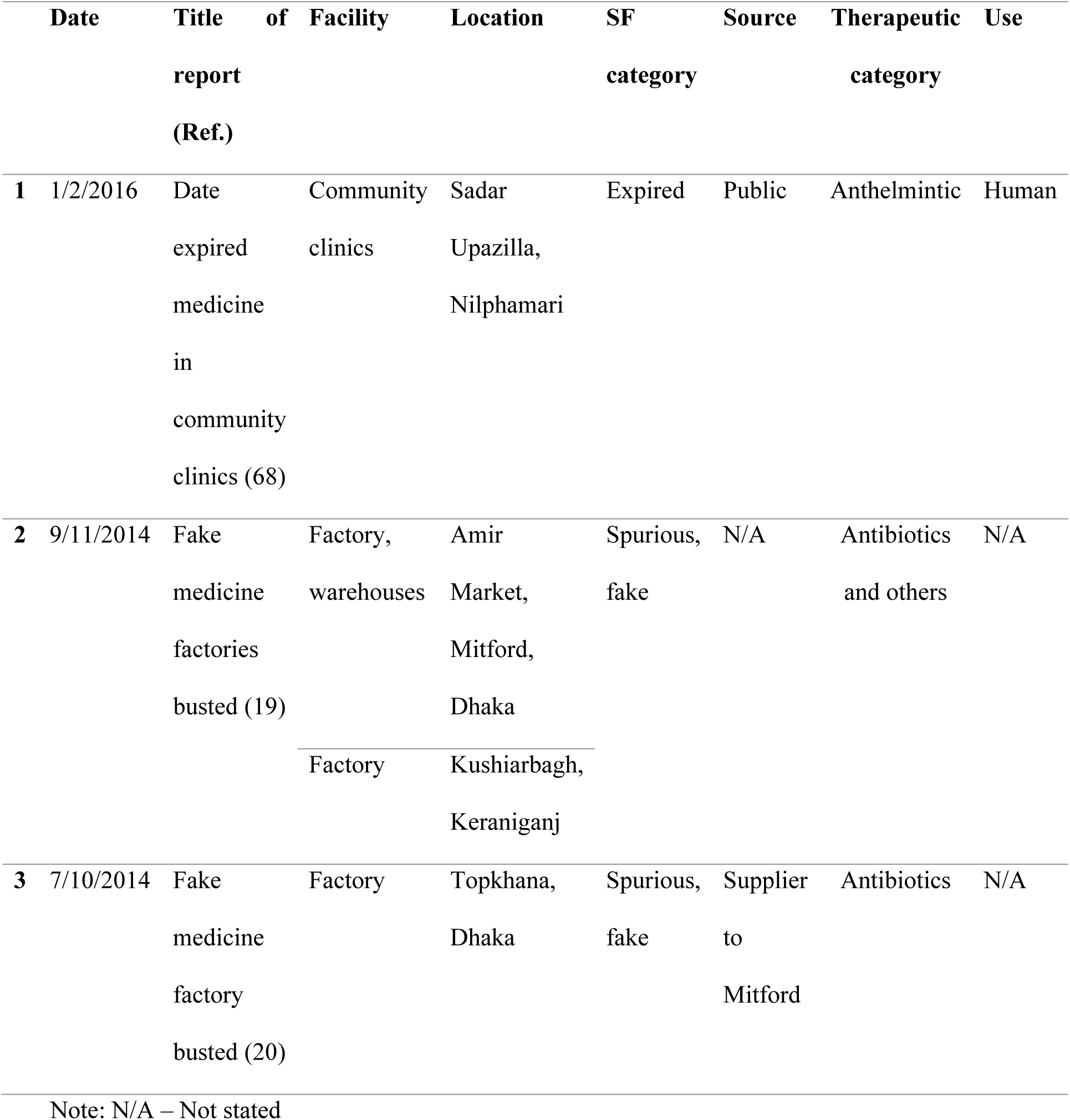
Newspaper reports of SF antimicrobials.

#### Reports on medicine quality

The search on Google Scholar retrieved six studies on the quality evaluation of antibiotics marketed in Bangladesh (Table 4). The EMBASE search retrieved three studies already in the Google Scholar search results. No relevant publications were identified via *Banglajol*.

**Table 4.**
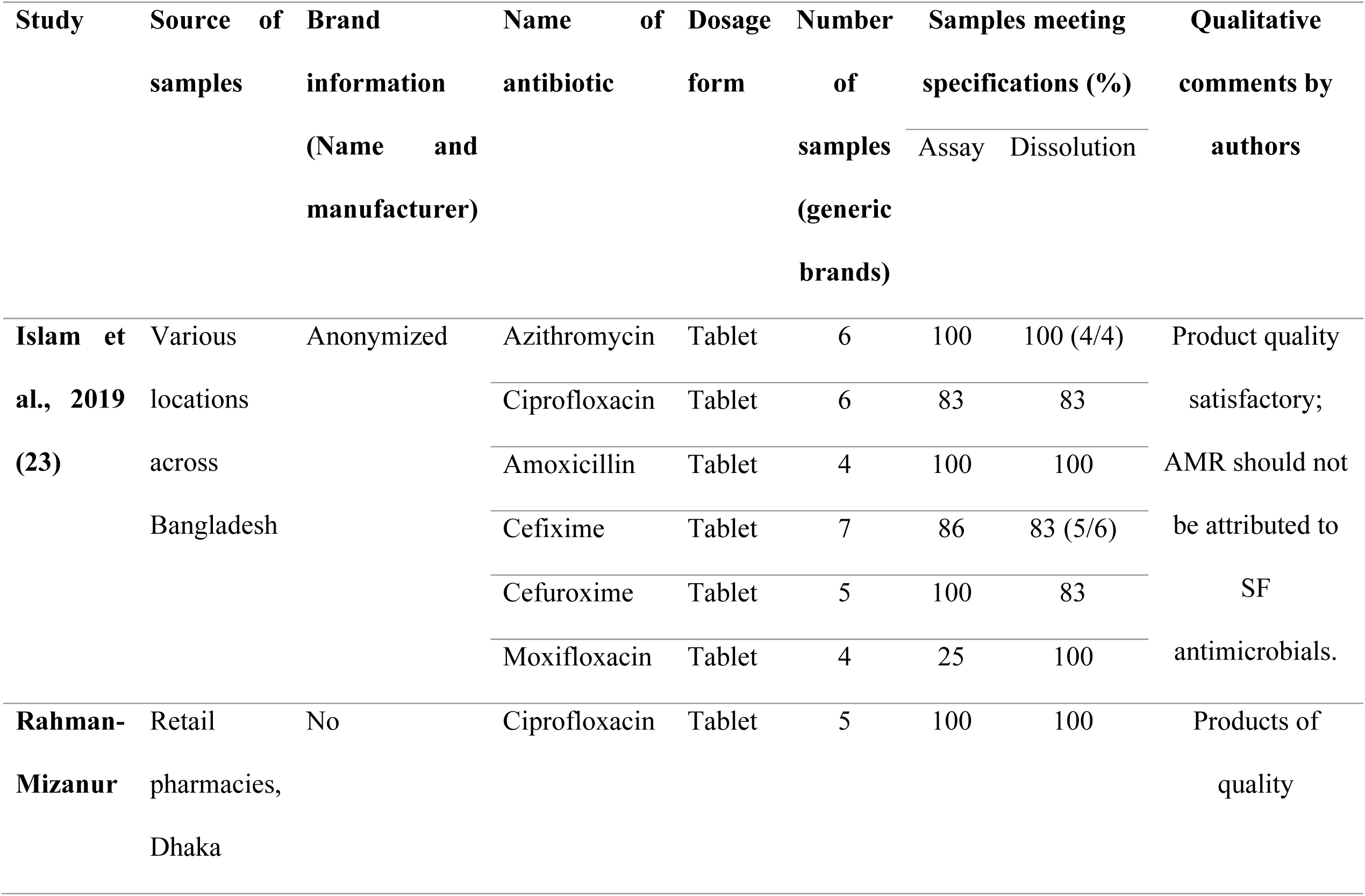
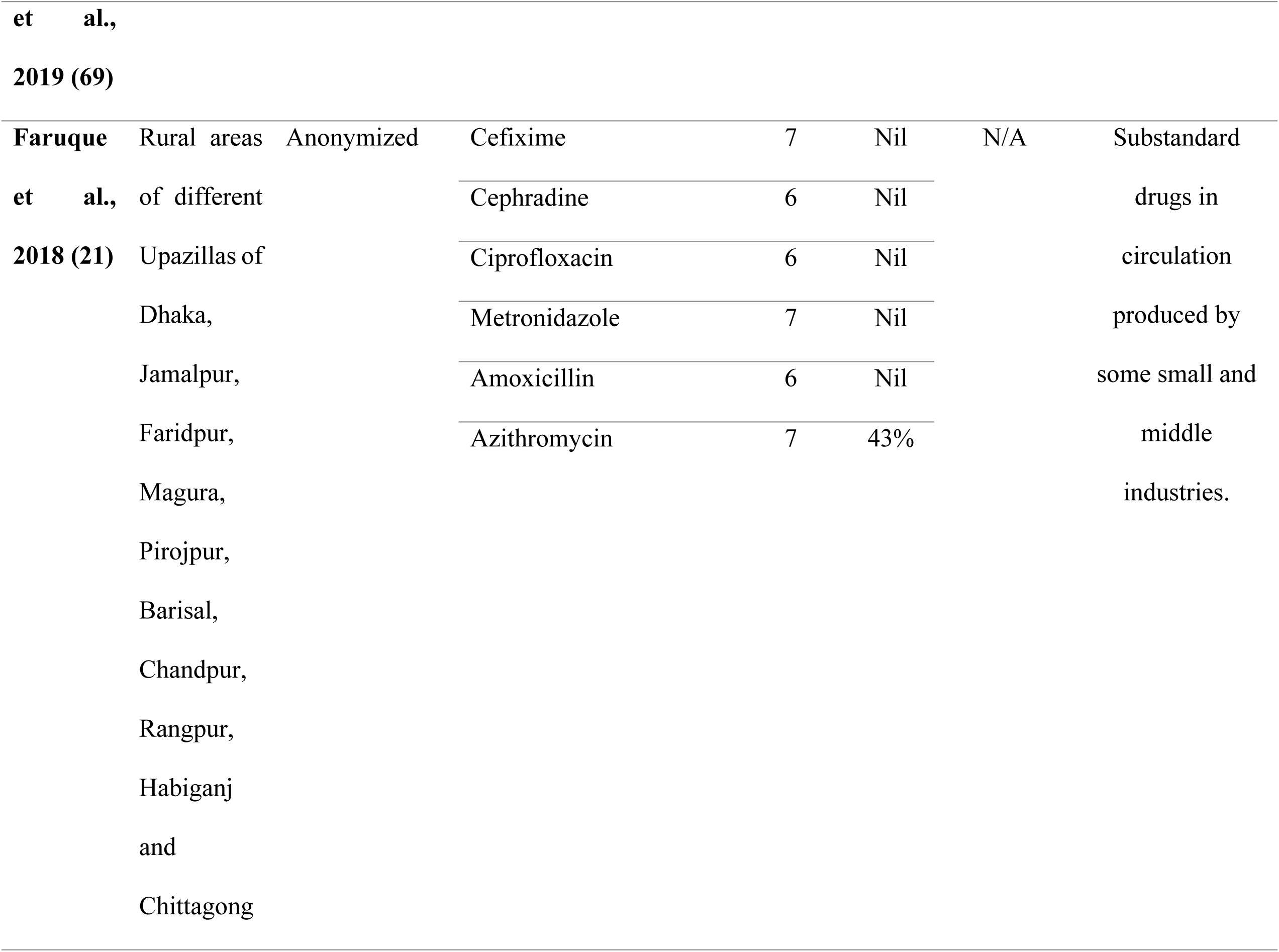
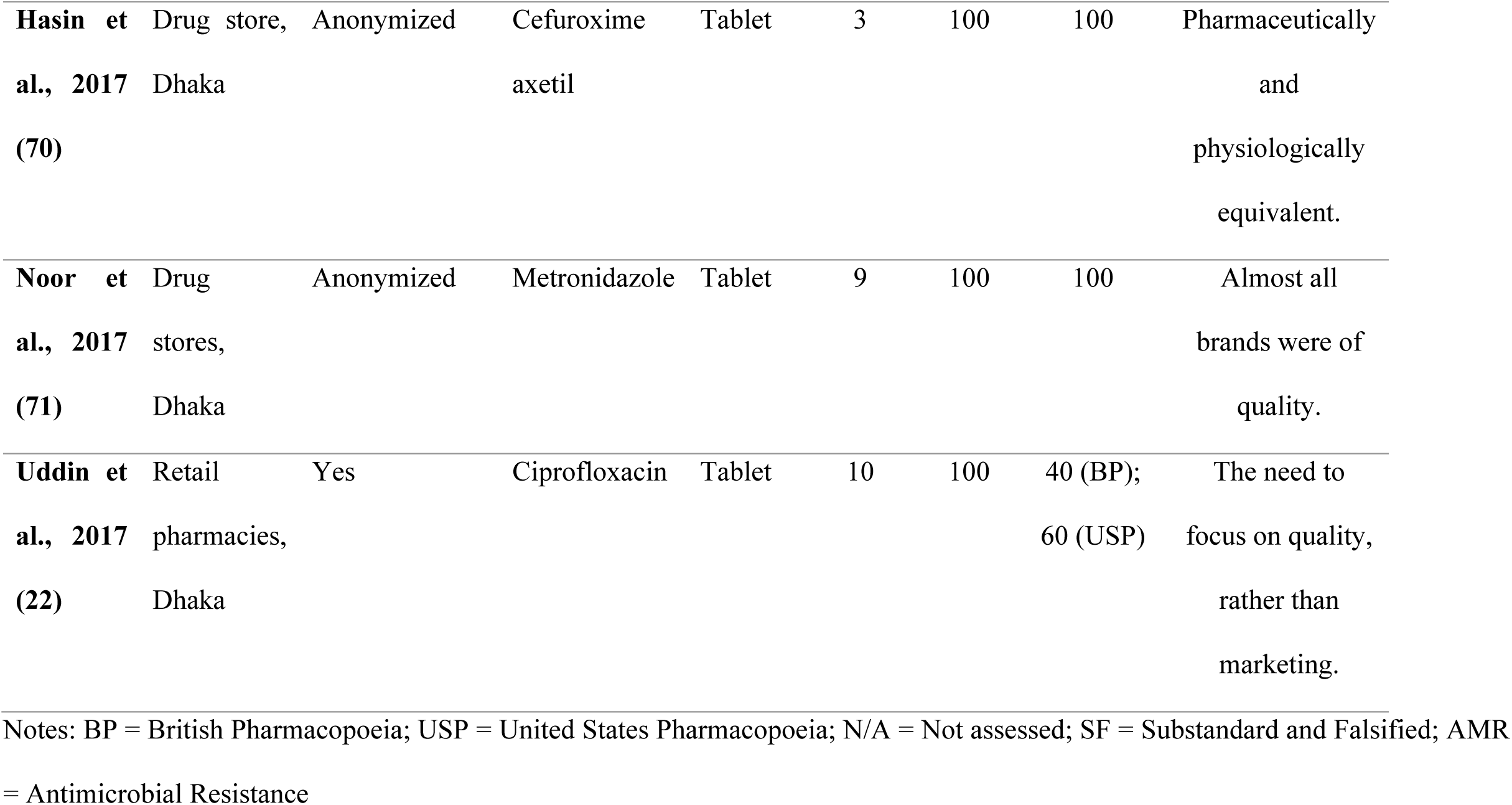
Reports of quality of antimicrobials in circulation in Bangladesh, 2017-2020.

Overall, the six studies tested between 1-6 different antibiotics from different manufacturers; the smallest study tested 3 different generics and the largest 39, where studies with larger samples found more out-of-specification results. Importantly, Faruque (2018) et al testing 39 samples of six antibiotics – cefixime, cephradine, ciprofloxacin, metronidazole, amoxicillin, and azithromycin – found all six evaluated antibiotics to be falsified, with no API content for five, and a low out-of-specification content for azithromycin. This study also suggests that SF medicines may be more prevalent in rural areas (21). Uddin (2017) found that 40% (4/10) different ciprofloxacin brands tested did not meet the dissolution specifications in the USP (or 60%, when evaluated by the British Pharmacopeia, BP) (22). In the study by Islam et al (2019), half (50%, 3/6) of the six different brands of antibiotics failed assay and/or dissolution (23).

Ciprofloxacin was the most commonly (67%, 4/6) evaluated antibiotic in all six studies. In three of these studies, the ciprofloxacin samples were substandard with out-of-specification dissolution profiles.

The link between AMR and SF medicine was investigated by Islam et al (2019), as one arm of a three-part study investigating resistance of selected bacteria to commonly used antibiotics and compliance. Even though half the samples failed one of the two tests for quality, the authors commented that products were generally of sufficient quality and that AMR should not be attributed to SF antimicrobials, but to the low rates (18%, n=300) of patients’ compliance.

#### Post-marketing surveillance

The MQDB and MQMG, as at the time of the study, did not hold records of any poor-quality antimicrobial from Bangladesh, apart from a WHO Alert on falsified oral cholera vaccine circulating in Bangladesh on MQMG.

We found no current reports from the DGDA’s website. In 2011, Bangladesh reported a failure rate of 0.04% of medicines assessed for quality as part of its pharmacovigilance activities (24).

## Discussion

In this study, we reviewed regulatory medicine quality assurance policies, media and post-marketing surveillance reports as well as antimicrobial quality studies in Bangladesh to provide an understanding of the local context and to suggest approaches to regulatory system strengthening. This assessment of the integrity of the antimicrobial supply chain in Bangladesh highlights the existence of regulations but a significant knowledge gap on the extent of SF antimicrobials. At the same time, it also highlights innovations and existing structures that could be leveraged to improve regulatory oversight in Bangladesh.

### Policy provisions, enforcement, and regulatory capacity challenges

This analysis shows that all iterations of the Bangladesh NDP recognize the need to ensure medicine quality throughout the supply chain and emphasize the need for manufacturers to fully comply with the WHO GMP guidelines. This is in consonance with other countries who have also incorporated these guidelines in their quality assurance mechanisms (25). The NDP also includes GDP guidelines for storage conditions as well as GPP guidelines.

In enforcing the drug laws, Bangladesh leverages other players including security and justice personnel. There is a standing Rapid Action Battalion (RAB) composed of DGDA personnel and security operatives assisted by a legislative vehicle, the Mobile Court, both to ensure implementation of existing regulations and to impose summary penalties, conducting post-marketing surveillance; similar to Nigeria (26,27).

However, there are regulatory capacity challenges. Overall, regulatory staff strength remains limited with 3,700 staff to oversee a supply chain with some estimated 200,000 distribution points and a product register of some 50,000 generics (28). In 2020, the DGDA maintains offices in only 47 out of the 64 districts of Bangladesh, indicative of structural challenges (18,29). These challenges are similar for other LMICs, including the world’s largest generics manufacturer, India (30,31).

### Market surveillance and medicine quality issues

Bangladesh, in addition to meeting 98% of its pharmaceutical needs, exports medicines to over a hundred countries (32,33). Its annual pharmaceutical exports is expected to cross the $1 billion mark in the next two years (34). Recently, the country produced its own remdesivir in a demonstration of its pharmaceutical manufacturing capacity (35).

Our analysis suggests some challenges with the antimicrobial supply chain in Bangladesh for the study period 2017-2020. Indeed, ever since the first report of poor-quality medicine in the 1980s of contaminated paracetamol, there has been recurrent reports in the newspapers of actions by RAB involving SF medicines – mostly falsified products or expired drugs (36–44).

The reports we described above found in the mass media are complemented by laboratory studies we identified from our literature review investigating the quality of antibiotics in circulation in Bangladesh and exploring the impact or implications. The findings are troubling considering that the evaluated antibiotics are among the top 10 most common licensed in Bangladesh (Unpublished results).

Despite the variability in choice of official reference – BP/USP – and methodological challenges with sample sizes, reporting styles, and authors perceptions/views of the implications of study findings, these studies point to the presence of SF antibiotics in the supply chain.

However, while the mass media and medicine quality studies suggest the persistence of SF medicines for human use, the extent of the challenge is not fully understood. In a dated report from 2011, Bangladesh reported a low SF prevalence of <1%. In 2019, the Consumer Protection Agency (CPA) survey estimated a high prevalence of expired medicines in pharmacies and medicine outlets in Bangladesh at 90% (n=600) (45).

### Developments, knowledge and implementation gaps, and suggestions for regulatory strengthening

These quality issues which persist in the presence of technically appropriate policies are syndromic of low regulatory capacity and call for improvements in regulatory systems. While there is evidence that this is already happening, for instance in technical analytical capacity, more is needed. In March 2020, the National Control Laboratory of the DGDA received WHO accreditation, two years after it attained the ISO/IEC 17025:2017 accreditation for laboratory testing and calibration (46). The NMRA has received assistance from the USP Pre-Qualification of Medicines (PQM) program to build capacity for the quality assurance of medicines manufactured and distributed in Bangladesh (47). In 2019, it received six Mini-Labs™, portable medicine quality testing devices capable of semi-quantitative analysis for rapid screening of medicine quality during post-marketing surveillance activities (48). However, there are other capacity challenges to medicine quality assurance of antimicrobials for human and veterinary use, apart from infrastructural, including human resources (29,49).

There is also need for a comprehensive evidence base on the prevalence and spatial distribution of SF antimicrobials to guide targeted and effective regulatory actions using this limited regulatory force. The government, or development partners, could commission a nation-wide quality audit of selected most consumed or marketed antimicrobials available for sale to the public. This would help to identify possible hot-spots for the manufacture, entry, distribution, or sale of SF antimicrobials for a risk-based approach to medicine quality assurance.

Bangladesh could address existing policy implementation gaps and further strengthen the integrity of the antibiotic supply chain by three approaches. Firstly, the use of existing community-based structures as a tool to strengthening medicine quality assurance. Interestingly, there are NGOs advocating for stricter actions on SF medicines, signifying public awareness and concern. For example, the Organization of Social and Environment Changes instituted a suit against the government in 2015 at the High Court questioning government’s apparent inaction to control the production of counterfeit medicines (50). These organizations may be leveraged to help with inspections of demand points, especially in hard-to-reach rural areas where SF medicines tend to proliferate, using a checklist created by the DGDA. This checklist which would be based on visual analysis by package inspection could include product licensing status and expiry date. Similar checklists have been proposed and studied (51,52). They could also help create community awareness on SF medicines. Interventions utilizing community-based approaches to improving health outcomes have been demonstrated in Bangladesh for reproductive health and TB management, for examples (53,54). Thus, this strategy could be a useful tool to explore in the context of pharmacovigilance and regulatory inspections.

Secondly, task-shifting approaches using designated academic institutions performing regular surveys on behalf of the DGDA may be another way to expand capacity. To strengthen output measures among several different research institutions, the WHO Guidelines on the conduct of Medicine Quality Assurance Survey could be adopted for uniformity in methodology and reporting (55). Mandating such independent analyses to be shared with the NMRA could provide synergies to improve the integrity of the supply chain in LMICs. While we found that the Bangladesh NDP mentions the use of third-partly medicine quality assessors, the extent to which this is utilized to complement efforts by the DGDA within Bangladesh is not known. Task-shifting has been widely used to address workforce capacity challenges in the health sector in many different countries, including high-income settings (56). There may be the need to extend pharmacovigilance also to the veterinary sector, using veterinarians and para veterinarians in the first instance to flag any suspected poor-quality veterinary medicines for follow-up action by the DGDA. The establishment of a separate veterinary medicines agency as is the case with Ethiopia may be another approach at capacity building (57).

Thirdly, scaling up existing technology platforms could lead to synergistic increases in overall regulatory capacity. Recently, as an extension of the national pharmacovigilance system, the DGDA launched a mobile Drug Verification application in 2019 for direct (voluntary) reporting by the public of adverse drug reactions, including of suspected counterfeit medicines (58,59).

Mandating all antibiotic manufacturers to include Mobile Authentication Service (MAS) codes on antibiotic packs would complement the Bangladeshi government initiative to enforce the sale of antibiotics only in packages, among others, as a means of promoting appropriate use. This track and trace approach is in use in different forms in countries and regions such as Dubai, Turkey, India and the European Union (60–65). Bangladesh under an innovative a2i project is leveraging information technology and a >80% mobile phone ownership in its population to provide services to the public. Track and trace technologies already exist in its significant fisheries/aquatic livestock sector (66). Thus, leveraging on a combination of end-user participation and technology could be one approach to improving supply chain integrity.

## Conclusions

This study found the existence of regulation and regulatory actions against poor-quality antimicrobials as well as news reports and peer-reviewed studies on poor-quality antimicrobials, but no current rigorous assessment of the extent of poor-quality antimicrobials. We suggest a multi-pronged diffuse approach to regulatory system strengthening comprising three strategies: community-based surveillance, task-shifting, and technology-enabled consumer participation.

These approaches could expand capacity to provide a more robust surveillance of the antimicrobial supply chain.

## Data Availability

All data produced in the present work are contained in the manuscript

## Ethical approval

Not required (as the study included only document and literature review and was not human subject related, there was no need for ethics).

## Acknowledgment

This work was made possible by support from the Boston University Social Innovation on Drug Resistance (SIDR) Postdoctoral Program to ESFO.

## Appendix

Market surveillance and quality issues: To map reports of SF medicines and market surveillance activities, we conducted a combined literature and database review as detailed:

### Literature review for reports on SF medicines: The search strategy is as described

Google was searched for “online Bangladesh English language newspapers”, retrieving 21 (72). Of these, that with the largest print circulation – the Daily Star – was purposively selected. The search term was “medicine”. Following a preliminary scan to identify local descriptors of poor-quality medicines, headlines/titles/captions and “abstracts”, or “one-liners” were then searched for any containing the following keywords: “fake”, “adulterated”, “substandard”, “closure”, “illegal”, “counterfeit”, “unwholesome”, “banned” or “expired”. Reports meeting these criteria were assessed for eligibility. Inclusion criteria were antimicrobial(s)/ antibiotic/ antihelminth(ic). Editorials and commentary pieces were excluded. Also excluded were reports on non-allopathic medicines including herbal and homeopathic medicines. Data was extracted in the form of the following variables: facility type (manufacturer, hospital or medicine outlet, trader, other) where SF medicines were found; location of facility; category of SF medicine; and whether medicine indicated for use in human or animals. Data were entered into an Excel spreadsheet for analysis.

### Reporting on quality testing

BANGLAJOL: The search on Banglajol was conducted using the search terms “medicine quality” and “drug quality”. Titles were screened for the same key words used in the literature search on Google above in addition to “quality”. Exclusion criteria were any study outside Bangladesh, studies on non-pharmaceutical substances including food and water.

EMBASE: The search on EMBASE was limited to publication year including 2016-2020 and filtered by drug. All antimicrobials used in human and animal health were selected.

GOOGLE SCHOLAR: The search on Google Scholar was limited from 2016-date; patents and citations were excluded before performing the search on Google Scholar. Search was conducted on May 18-19, 2020. The search term was “drug quality Bangladesh”. Retrieved articles were screened for relevance. Titles and abstracts containing “quality” AND/OR “Bangladesh” and describing a laboratory analysis were collated into a reference manager software. Laboratory analyses were identified by any, or combinations of, the following terms: “*in vitro*”, “HPLC”, “dissolution”, “potency”, “assay”, “pharmacopeial”, “evaluation”, “analysis”, or “release”.

### Articles were then assessed for eligibility for evaluation using the following criteria

∘ Inclusion: Only articles describing a laboratory analysis of an antibiotic were evaluated.
∘ Exclusions: Studies conducted outside of Bangladesh studies; studies on method development without comments on medicine quality; quality studies on diagnostic or clinical care; articles describing prevalence or incidence without primary data on compendial analysis; studies on other therapeutics groups not including antibiotics. Data extracted included: Facility type and location from which samples were collected; name of samples, where a sample was one antimicrobial agent identified by the International Non-proprietary Name (INN), number of samples (brands), official tests, and proportions of samples passing the tests. To ensure comparability, data was collected only for two official tests – potency (assay) and dissolution. The qualitative comments of authors in the discussion section of the articles were noted to gauge overall perceptions. Any article linking poor-quality to AMR was noted.

